# Deep learning of point-of-care lung ultrasound and clinical data for pulmonary tuberculosis triage in Uganda

**DOI:** 10.64898/2026.11.14.26368125

**Authors:** Akanksha Sharma, Lydia Nakiyingi, Elena Jovanovska Janeva, Keith Guevarra, Wycliff Nate, Eva Nabawanuka, Raphael Shankman, Rohan Dhamdere, Guadalupe Astorga, Moses Joloba, Jerrold Ellner, Anant Madabhushi, Yingda L. Xie

## Abstract

**Rationale:** Point-of-care lung ultrasound is portable and radiation-free and could extend tuberculosis screening to settings without chest radiography, but no standardized criteria exist for interpreting it for tuberculosis.

**Objectives:** To develop and internally evaluate deep learning models that classify pulmonary tuberculosis from lung ultrasound images, alone and combined with routine clinical variables, and to derive an abbreviated scanning protocol.

**Methods:** In a prospective study in Kampala, Uganda (2023–2025), we enrolled people aged 13 years or older with microbiologically confirmed pulmonary tuberculosis, their household contacts, and symptomatic people with other respiratory conditions. One still frame from each of 12 lung ultrasound zones was analyzed with an attention-based multiple-instance learning network. We compared clinical-only (random forest), ultrasound-only, and combined models and derived a 6-zone protocol from training-set attention weights. Participants were allocated to training (n = 223), validation (n = 20), and held-out test (n = 73) sets.

**Results:** Of 318 enrolled participants, 316 were analyzed (118 with and 198 without tuberculosis). In the test set, areas under the receiver operating characteristic curve were 0.88 (95% confidence interval, 0.80–0.95) for the clinical-only model; 0.93 (0.86–0.97) and 0.95 (0.89–0.99) for the 12-zone ultrasound-only and combined models; and 0.91 (0.83–0.97) and 0.93 (0.86–0.99) for the corresponding 6-zone models. At 90% sensitivity, specificity was 82.2% (34.9–97.8) for the 12-zone combined model and 91.1% (25.0–98.0) for the 6-zone combined model. Point estimates for all 12-zone models and the 6-zone combined model met the 2025 World Health Organization minimum accuracy targets for a high-sensitivity screening test (≥90% sensitivity, ≥60% specificity).

**Conclusion:** Lung ultrasound analyzed with attention-based deep learning and combined with clinical variables showed high internal discrimination for pulmonary tuberculosis, and a simplified 6-zone protocol retained most of this performance. External validation in intended-use screening populations is needed.

## I. INTRODUCTION

Tuberculosis (TB) remains a leading driver of morbidity and mortality in low- and middle-income countries. TB burden is concentrated among people living with HIV and undernutrition, and diagnostic gaps are widest in settings without consistent access to radiography, microbiological testing, or trained radiologists (1). Chest radiography is widely used for TB screening and triage (2). Its deployment, however, is constrained by equipment cost, maintenance, radiation exposure, and infrastructure (3).

Point-of-care lung ultrasound (POCUS; LUS) is portable, inexpensive, and radiation-free, and can be acquired at the bedside by trained non-radiologist operators (3,4). However, sonographic findings in pulmonary TB, such as subpleural nodules, consolidation, and B-lines, are not specific to TB, and no standardized criteria exist for interpreting them (3). Emerging proof-of-concept studies to date have evaluated artificial intelligence (AI)-assisted LUS for TB. In a prospective study of 504 symptomatic adults in urban Benin, an ensemble of deep learning models achieved an area under the receiver operating characteristic curve (AUC) of 0.93, with 93% sensitivity and 81% specificity (5). A second model, a convolutional neural network with long short-term memory layers trained on video clips from 497 Zambian participants to detect radiographer-labeled lung abnormalities, achieved AUCs of 0.89–0.92 against a composite TB reference standard (6).

We developed SCAN-TB (Sonographic and Clinical AI-guided Navigator for TB triage) which builds on these advances in three ways. First, we trained models directly on participant-level microbiological TB status using attention-based multiple-instance learning, which treats all lung-zone images from a participant as one diagnostic unit and estimates each zone’s contribution. Second, we used zone-level attention to derive a 6-zone protocol as a shorter alternative to a full 12-zone acquisition. Third, we hypothesized that combining lung ultrasound with routinely collected clinical variables would discriminate TB better than either source alone. Here we report model development and internal evaluation in an urban Ugandan cohort.

## II. METHODS

### Study design and participants

We conducted a prospective observational sub-study within the Bacterial a d Host Determinants in TB Transmission project (TBRU/Halting TB) in Kampala, Uganda. POCUS-TB participants were enrolled between July 2023 and March 2025 at TBRU/Halting TB enrollment sites and Kampala Capital City Authority health facilities. The study is reported according to the TRIPOD+AI (7) and study workflow is summarized in Figure 1.

**Figure 1.**
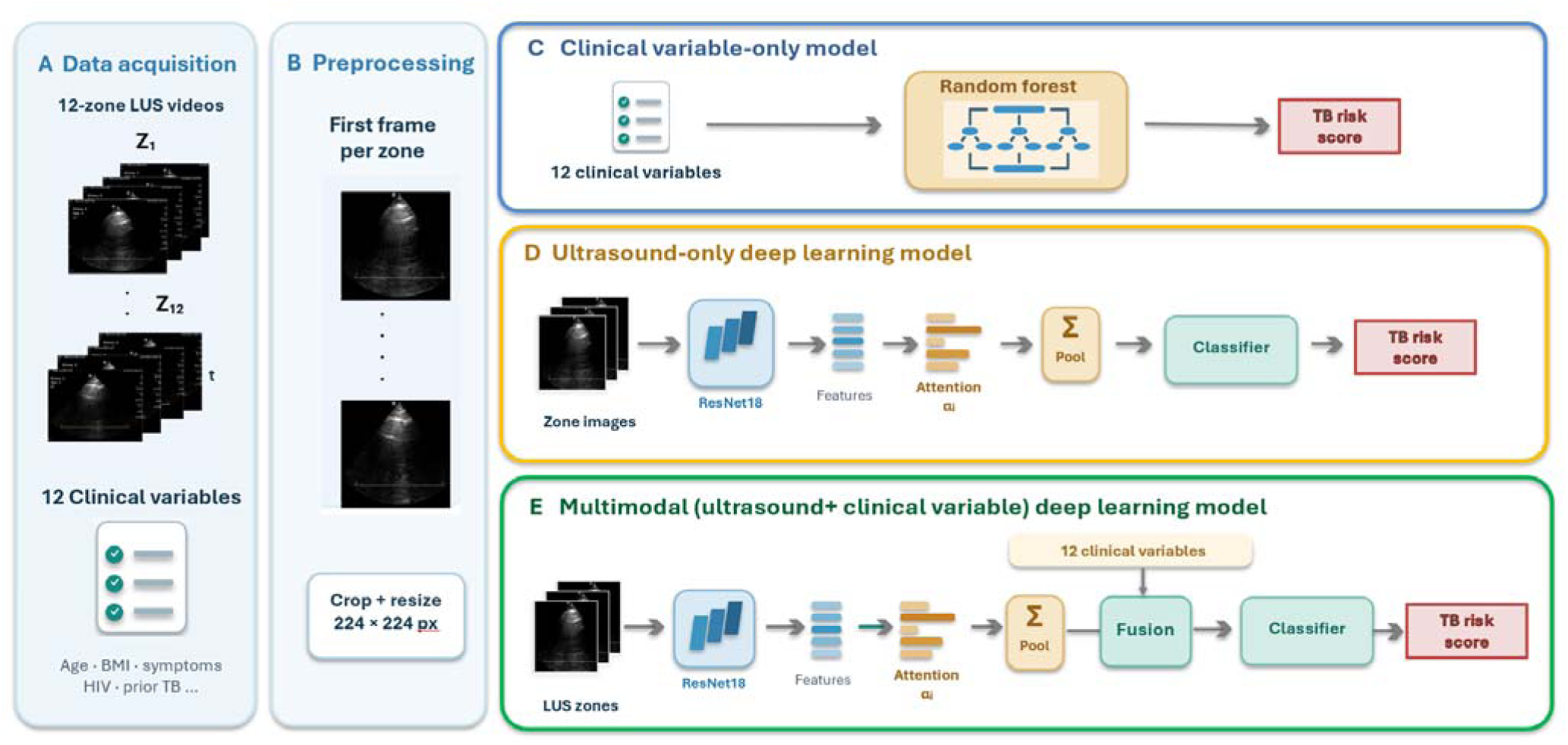
Study overview. (A) Acquisition of lung ultrasound clips from 12 zones and of clinical variables. B) Preprocessing: selection of the first frame from each zone, cropping, and removal of on-screen text. (C) Clinical variable-only random forest model. (D) Ultrasound-only model: ResNet18 encoder with attention-based multiple-instance learning pooling of zone embeddings. (E) Combined model: concatenation of clinical variables with the pooled image embedding (intermediate, feature-level fusion). Alt text: Schematic of the study workflow, showing collection of lung ultrasound clips and clinical variables, selection of one frame per lung zone, and three models: clinical variable-only random forest, ultrasound-only deep learning, and combined ultrasound and clinical-variable deep learning.

Eligible participants were individuals aged 13 years or older who (i) had laboratory-confirmed pulmonary TB, (ii) were household contacts (HHCs) of an individual with confirmed pulmonary TB, or (iii) were evaluated for TB and diagnosed with another respiratory condition. Index cases were enrolled consecutively from parent-study participants who met eligibility criteria. Individuals who had received >15 days of TB treatment in the 30 days before enrollment were excluded. This design was intended to represent a broad spectrum of disease: early TB among recently-exposed household contacts, symptomatic and often more advanced TB among index cases, and TB-negative individuals ranging from healthy contacts to symptomatic patients with competing respiratory diagnoses.

### Sample size

The protocol targeted approximately 235 TB+ and 300 TB− participants, with 20% held out for evaluation. This would provide at least 80% power to show sensitivity ≥90% (against ≤70%) and specificity ≥70% (against ≤50%). Enrollment closed at the end of the funded study period with 118 TB+ and 198 TB− participants. The held-out test set (28 TB+, 45 TB−) met the planned requirement for specificity but not for sensitivity, thus sensitivity estimates are less precise than planned.

### Ethics statement

The study was approved by Mulago Hospital Research Ethics Committee (2349) and the Rutgers University Institutional Review Board (Pro2020003026). It was conducted in accordance with the Declaration of Helsinki. Adult participants provided written informed consent. Participants aged 13–17 years provided written assent, with written consent from a parent or guardian. Patients and the public were not involved in the design, conduct, reporting, or dissemination of this research.

### Reference standard and participant classification

Pulmonary TB status was based on microbiologic detection of *Mycobacterium tuberculosis* in sputum. Index cases and household contacts underwent both Xpert MTB/RIF Ultra (Ultra) and Mycobacteria Growth Indicator Tube (MGIT) culture. Participants recruited from clinic patients undergoing TB evaluation who were diagnosed with another respiratory condition underwent Ultra. Participants with an Ultra- or culture-positive result were classified as TB-positive (TB+). TB-negative participants were classified by clinician diagnosis as having no respiratory condition (NRC) or another, non-TB respiratory condition (ORC). For modeling, NRC and ORC participants were combined as TB-negative (TB−).

### Lung ultrasound acquisition

All participants underwent a standardized LUS examination with a Philips Lumify phased-array probe (Philips Healthcare) connected to an Android tablet. Six-second clips (30 frames/s, 768 × 1024 pixels) were recorded from 12 zones covering the anterior, lateral, and posterior chest bilaterally (R1–R6 and L1–L6; Figure S1). All examinations were performed by one of two study technicians who had completed in-person training and competency certification in acquisition and preliminary interpretation. Technicians were blinded to laboratory results, clinical diagnosis and treatment status, and chest radiograph findings. Participants with no recorded zones were excluded (Figure 2).

**Figure 2:**
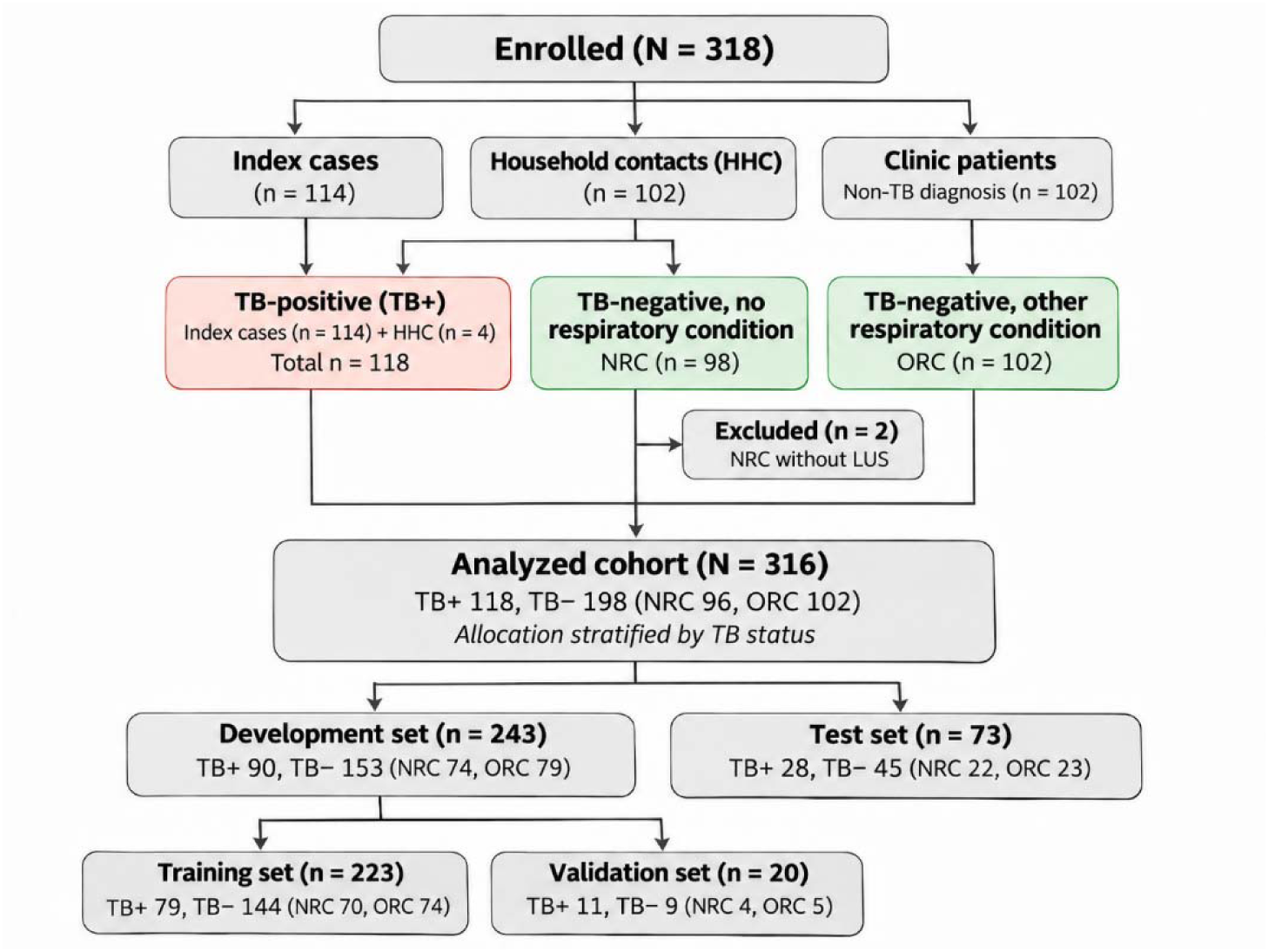
Participant flow, exclusions, and dataset partitioning for model development and evaluation. Household contacts with *Mycobacterium tuberculosis* detected in sputum were classified as TB-positive. Allocation to the development and test sets was stratified by TB status, and the validation subset was drawn from the development set. HHC = household contact; LUS = lung ultrasound; NRC = TB-negative, no respiratory condition; ORC = TB-negative, other respiratory condition; TB+ = Ultra- or culture-positive pulmonary TB. Alt text: Flow diagram of 318 enrolled participants grouped as index cases, household contacts, and clinic patients, then classified as tuberculosis-positive, no respiratory condition, or other respiratory condition. Two participants were excluded, and the remaining 316 were divided into training, validation, and test sets.

### Data partitioning

Participants were randomly allocated at the participant level to training (n = 223, 70.6%), validation (n = 20, 6.3%), and held-out test (n = 73, 23.1%) sets. Allocation to the development (training and validation) and test sets was stratified by TB status, giving similar TB+ proportions of 37.0% (90/243) and 38.4% (28/73), respectively. The validation set was used for early stopping and checkpoint selection. The checkpoint with a validation loss improvement greater than 10^-3^ was saved. All performance estimates are from the test set.

### Clinical variables

Twelve clinical variables associated with TB risk were recorded at enrollment on standardized case report forms: age (8), sex (9), history of prior TB (10), HIV status (11), diabetes (12), smoking history (years) (13), cough duration (weeks) (14), weight loss duration (weeks)(14), participant general appearance, temperature (14), body mass index (BMI) (15–17), and number of household members (18). Variable definitions and coding are given in Table S1. A random forest classifier with default scikit-learn hyperparameters was trained on the training set. Variables were z-score normalized using training-set means and standard deviation, which were then applied to the validation and test sets. Variable contributions were ranked by mean absolute Shapley additive explanation (SHAP) values (19).

### Deep learning ultrasound and combined models

Every fourth frame was retained. Frames were automatically cropped to remove on-screen text and borders and resized to 224 × 224 pixels. The first frame of each zone was selected a priori as a single, reproducible frame-selection rule, and the middle frame was used in a sensitivity analysis. Each zone image was encoded with a ResNet18 network (20). Zone embeddings were aggregated per participant with attention-based multiple-instance learning pooling (21), with attention weights softmax-normalized across each participant’s zones to sum to 1. The ultrasound-only model classified the pooled embedding directly. In the combined model, the 12 normalized clinical variables were concatenated with the pooled embedding before the classification layers (intermediate, feature-level fusion), and the network was trained end to end. Models classified TB+ versus TB− and were trained on the observed class distribution with unweighted cross-entropy loss, the AdamW optimizer, and early stopping on validation loss; no resampling or class weighting was applied. Detailed network parameters and architectural specifications are provided in Supplementary Figure S2. Parameters related to model training are listed in Supplementary Tables S3 and S4, and training and validation loss and AUC curves are shown in Supplementary Figure S3.

To interpret the model’s decision-making process, integrated-gradient maps (22) were generated to highlight image regions that contributed most strongly to the model output. These maps were evaluated alongside the corresponding zone-level attention scores, characterizing contributions both within and between zones. Additionally, Uniform Manifold Approximation and Projection (UMAP) (23) projected participant embeddings into two dimensions to visualize separation of participant groups.

### Selection of the 6-zone protocol

To explore an abbreviated protocol from the 12-zone examination, we prioritized zones using the attention weights learned by the 12-zone ultrasound-only model. For each zone, we computed the mean attention weight across the TB+ participants in the training set (class-specific mean attention). Zones were ranked by this value, and the six highest-ranked zones were retained (Table S6). The 6-zone protocol was evaluated retrospectively using these zones from the 12-zone examinations.

### Statistical analysis

All models performed binary classification of TB+ versus TB− (NRC and ORC combined). Performance was evaluated in the test set using the AUC and the F1 score at a probability threshold of 0.5. We also assessed three operating points. We also assessed three operating points. Sensitivity at 70% specificity reflects the 2014 WHO triage target product profile minimum of 90% sensitivity and 70% specificity (24). Sensitivity at 60% specificity and specificity at 90% sensitivity reflect the 2025 WHO minimum for a high-sensitivity screening test (25). The classification model was built to perform 2-class classification, i.e. TB+ vs TB-(NRC and ORC). Sensitivity and specificity at these operating points were estimated by linear interpolation of the empirical test-set ROC curve. These analyses were intended to characterize model performance relative to WHO benchmarks and do not establish fixed decision thresholds for clinical deployment. Point estimates were computed on the full test set. 95% confidence intervals (CIs) for the AUC, sensitivity, and specificity were estimated by participant-level percentile bootstrap with 1,000 resamples. AUCs were compared pairwise with DeLong’s method (26), and AUC differences are reported with CIs and unadjusted P values. Accuracy by sex, HIV status, and prior TB was examined as an exploratory analysis. Performance was not compared across enrollment sites because of the small number of test participants per site. Baseline characteristics are reported descriptively, with comparisons in Table S5. Analyses were performed in Python 3.9, using PyTorch 2.3.1 for the deep learning models and scikit-learn 1.6.1 for the random forest model and bootstrap estimates.

## III. RESULTS

### Baseline characteristics

Of 318 enrolled participants, 114 were index cases, 102 were HHCs, and 102 were ORCs recruited from clinic patients. ORC diagnoses included other pneumonia (bronchopneumonia, lobar, or interstitial; 41.2%), upper respiratory tract infections (influenza, common cold, pharyngitis, tonsillitis, or rhinosinusitis; 24.5%), bronchitis (10.8%), allergic or vasomotor rhinitis (8.8%), obstructive airway disease (8.8%), and other conditions (pleural effusion, pulmonary edema, interstitial lung disease, and atelectasis; 5.9%). On microbiologic evaluation, 4 HHCs were classified as TB+ and none were classified as ORC. Two NRC participants were excluded because no LUS zones were recorded. The analyzed cohort therefore comprised 316 participants: 118 TB+ (37.3%), 96 NRC (30.4%), and 102 ORC (32.3%) (Figure 2). The training set included 79 TB+, 70 NRC, and 74 ORC participants; the validation set 11, 4, and 5; and the test set 28, 22, and 23. Compared with TB-participants, TB+ participants had lower median BMI (19.4 vs 22.4 kg/m²), longer cough duration (12 vs 2 weeks), and longer weight-loss duration (4 vs 0 weeks). They were less often rated not ill (6.8% vs 50.5%) and more often reported current or former smoking (20.3% vs 10.6%). Age, sex, HIV prevalence, prior TB, and diabetes did not differ between groups (Table 1; S2 by training/validation/test cohorts). Among TB+ participants, 111 (94.1%) were Ultra-positive, 96 (81.4%) were culture-positive, and 90 (76.3%) were smear-positive.

**Table 1.** Participant characteristics by tuberculosis status.

|  | All (n=316) | TB+ (n=118) | TB- (n=198) | p-value |
|---|---|---|---|---|
| Male, N (%) | 178 (56) | 72 (61) | 106 (54) | 0.195 |
| Age (years); median (IQR) | 32 (25–43) | 32 (25–40) | 32 (24–46) | 0.648 |
| BMI (kg/m²); median (IQR) | 20.6<br>(18.9–24.2) | 19.4<br>(17.8–20.8) | 22.4<br>(19.8–26.0) | <0.001 |
| Temperature (°C); median (IQR) | 36.5<br>(36.2–36.8) | 36.5<br>(36.2–36.8) | 36.4<br>(36.1–36.7) | 0.091 |
| Prior TB history, N (%) | 17 (5) | 5 (4) | 12 (6) | 0.487 |
| Diabetes, N (%) | 2 (1) | 1 (1) | 1 (1) | 1.000 |
| HIV positive, n (%) | 64 (20) | 23 (19) | 41 (21) | 0.795 |
| Current or former smoking, n (%)<sup>c</sup> | 45 (14) | 24 (20.3) | 21 (10.6) | 0.017 |
| Former | 24 (8) | 12 (10) | 12 (6) |  |
| Current | 21 (7) | 12 (10) | 9 (5) |  |
| Cough duration (weeks); median (IQR) | 4 (1–12) | 12 (4–16) | 2 (0–4) | <0.001 |
| Weight loss duration (weeks); median (IQR) | 0 (0–4) | 4 (3–8) | 0 (0–0) | <0.001 |
| General appearance,<br>Not ill/ Mildly ill / Moderately ill, n (%)† | 108 (34) /<br>150 (47) /<br>58 (18) | 8 (7) /<br>72 (61) /<br>38 (32) | 100 (51) /<br>78 (39) /<br>20 (10) | <0.001 |
| Known TB contact, N (%) | 41 (14) | 23 (19) | 18 (11)<sup>#</sup> | 0.053 |
| Household members, median (IQR)‡ | 4 (3–5) | 4 (3–5) | 4 (3–5) |  |
| Laboratory-confirmed (Ultra or culture positive), No./total (%) | — | 118/118<br>(100.0) | — | — |
| Xpert Ultra positive, No./total (%) | — | 111/118 (94.1) | — | — |
| High / medium / low / very low, n |  | 39 / 34 / 32 / 6 |  |  |
| Culture positive (MGIT or LJ), No./total (%) | — | 96/118 (81.4) | — | — |
| Smear positive, No./total (%) | — | 90/118 (76.3) | — | — |
| Clinical diagnosis only, No./total (%) | — | 0/118 (0.0) | — | — |

BMI = body mass index; IQR = interquartile range; TB = tuberculosis; TB+ = Xpert MTB/RIF Ultra- or culture-positive pulmonary TB; TB− = participants with no respiratory condition (n = 96) and with another respiratory condition (n = 102) combined. No data were missing except as noted. \*Six participants aged 13–17 years with smoking recorded as not applicable were classified as never having smoked; the models used smoking duration in years. †No participant was rated gravely ill. <sup>#</sup>TB contact variables were not collected for 32 ORC participants; denominator includes 166 TB-. ‡Top-coded at 8 (n = 8).

### Clinical variable only model

The clinical-only random forest model achieved an AUC of 0.88 (95% CI, 0.80– 0.95) (Table 2). The variables with the largest SHAP values were weight-loss duration, cough duration, BMI, general appearance, and number of household members (Figure 3). Diabetes, present in two participants, contributed least.

**Figure 3:**
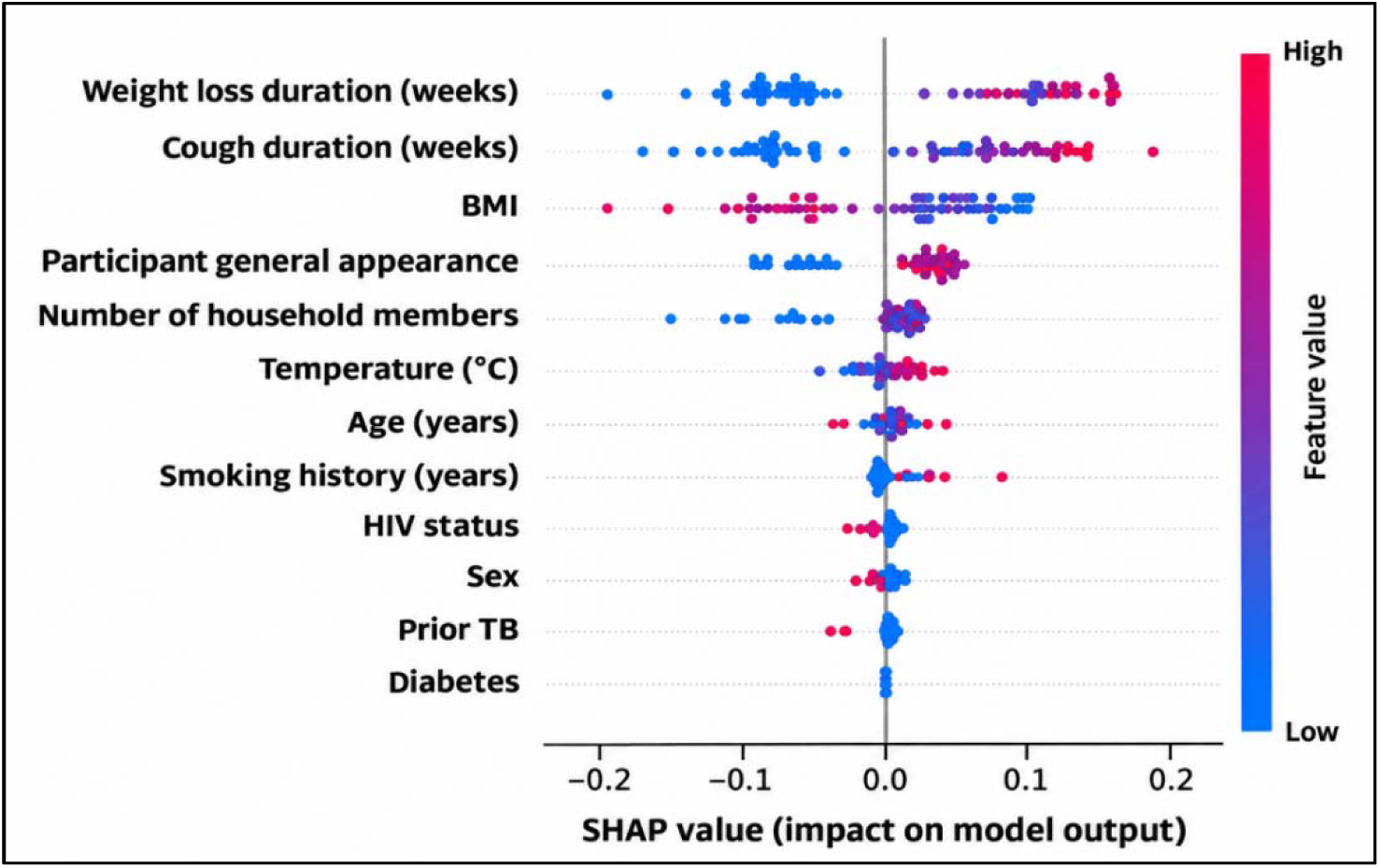
Contribution of clinical variables to the clinical-only model. SHAP summary plot in which each point is one participant in the test set. Positive SHAP values favor a TB-positive prediction, color indicates the observed variable value, and variables are ordered by mean absolute SHAP value. Color code for Sex: Female-Red, Male-Blue; Color code for General appearance, Diabetes, Prior TB, HIV status, Yes-Red, No-Blue. Alt text: Dot plot ranking 12 clinical variables by their contribution to the clinical-only model’s predictions. Weight-loss duration, cough duration, and body mass index contributed most, with longer symptom duration and lower body mass index favoring a tuberculosis prediction.

**Table 2:** Test-set performance of clinical-only, ultrasound-only, and combined models (n = 73; 28 TB+, 45 TB−). SN = sensitivity. SP = specificity

| **Zones** | **Model** | **AUC<br>(95%CI)** | **F1** | **SN @ 70% SP<br>% (95% CI)** | **SN @ 60% SP<br>% (95% CI)** | **SP @ 90% SN<br>% (95% CI)** |
|---|---|---|---|---|---|---|
|  | Clinical-only | 0.88<br>(0.80–0.95) | 0.74 | 89.3<br>(70.6–100) | 100<br>(81.2–100) | 66.2<br>(52.4–86.7) |
| 12 | Ultrasound-only | 0.93<br>(0.86–0.97) | 0.79 | 96.4<br>(77.8–100) | 96.4<br>(85.7–100) | 73.3<br>(41.9–91.3) |
|  | Combined | 0.95<br>(0.89–0.99) | 0.87 | 96.4<br>(87.5–100) | 96.4<br>(88.0–100) | 82.2<br>(34.9–97.8) |
| 6 | Ultrasound-only | 0.91<br>(0.83–0.97) | 0.79 | 85.7<br>(71.1–96.8) | 89.3<br>(73.7–100) | 57.8<br>(32.7–95.4) |
|  | Combined | 0.93<br>(0.86–0.99) | 0.86 | 92.9<br>(82.1–100) | 92.9<br>(81.0–100) | 91.1<br>(25.0–98.0) |

Definition of abbreviations: AUC = area under the receiver operating characteristic curve; CI = confidence interval; F1 = harmonic mean of precision and recall at a probability threshold of 0.5; SN = sensitivity; SP = specificity. Sensitivity and specificity at fixed operating points were estimated by linear interpolation of the test-set ROC curve; CIs are from 1,000 bootstrap resamples.

### Selected zones

The six highest-ranked zones by mean attention were R5 (0.109), L5 (0.096), L3 (0.088), R1 (0.083), R3 (0.082), and L1 (0.081), comprising the right and left anterior superior, lateral superior, and posterior superior zones (Table S6). Uniform attention across 12 zones would give 0.083. The seventh- and eighth-ranked zones, R6 and L4, each had a mean attention of 0.080.

### Ultrasound-only and combined models

In the 12-zone protocol, AUCs were 0.93 (95% CI, 0.86–0.97) for the ultrasound-only model and 0.95 (0.89–0.99) for the combined model. In the 6-zone protocol, they were 0.91 (0.83–0.97) and 0.93 (0.86–0.99), respectively (Table 2; Figure 4). At 90% sensitivity, specificity was 73.3% and 82.2% for the 12-zone models and 57.8% and 91.1% for the 6-zone models. The 6-zone ultrasound-only model fell just short, with 89.3% sensitivity at 60% specificity and 57.8% specificity at 90% sensitivity. Only the two 12-zone models and the 6-zone combined model also met the 2014 WHO triage target (≥90% sensitivity at ≥70% specificity). With the middle frame, AUCs were 0.85 (0.74–0.94) for the 12-zone ultrasound-only model and 0.90 (0.82–0.97) for the 12-zone combined model (Table S7).

**Figure 4:**
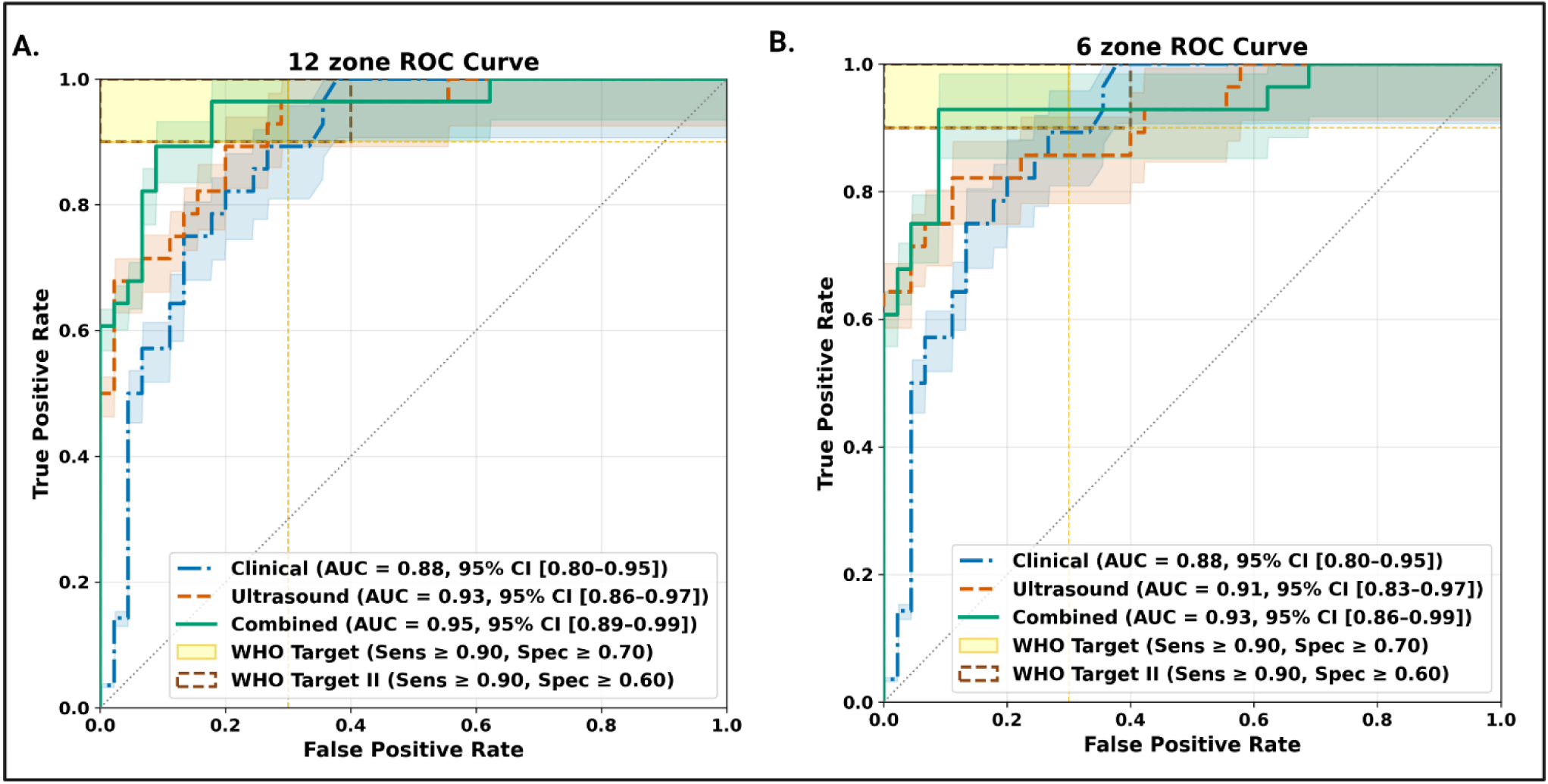
Receiver operating characteristic curves in the test set. Panel A shows the 12-zone protocol and panel B the 6-zone protocol, each with the clinical-only model for reference. Markers indicate the operating points at 70% and 60% specificity and at 90% sensitivity. Alt text: Two panels of receiver operating characteristic curves comparing clinical-only, ultrasound-only, and combined models for the 12-zone and 6-zone protocols. In both panels, the combined model’s curve is highest and reaches the shaded World Health Organization target regions.

### Exploratory subgroup analysis

Model performance was too few to assess, with no statistically significant differences in classification accuracy detected (Fisher’s Exact Test: HIV status OR=1.06, p=1.00; sex OR=0.9569, p=1.00; prior TB p=1.00; Figure S4). Accuracy was used in place of AUC given the limited subgroup sample sizes (n=3 prior TB) and (n=20 HIV-positive) in the test set, which precluded stable ROC curve estimation.

### Model interpretation

Figure 5 shows attention weights and integrated-gradient maps from the 6-zone ultrasound-only model for two TB+ participants with several pathologic features. In the first participant, subpleural nodules were present in L1, L3, and L5. R1 showed a shred sign, R3 showed subpleural nodules with B-lines, and R5 showed B-lines; attention was highest for L1. In the second participant, subpleural nodules or consolidations were present in R1 and L5, and these zones received the highest attention. Attribution was imperfect, with some emphasis on less relevant regions, including deeper lung tissue and artifacts, while some visible abnormalities received little attribution. In UMAP projections of the 6-zone models (Figure S5), TB+ participants overlapped with NRC and ORC participants in the clinical-only model. They formed a more distinct cluster in the ultrasound-only model, and the combined model showed the least overlap among the three groups, with tighter within-cluster cohesion and greater between-cluster margins.

**Figure 5:**
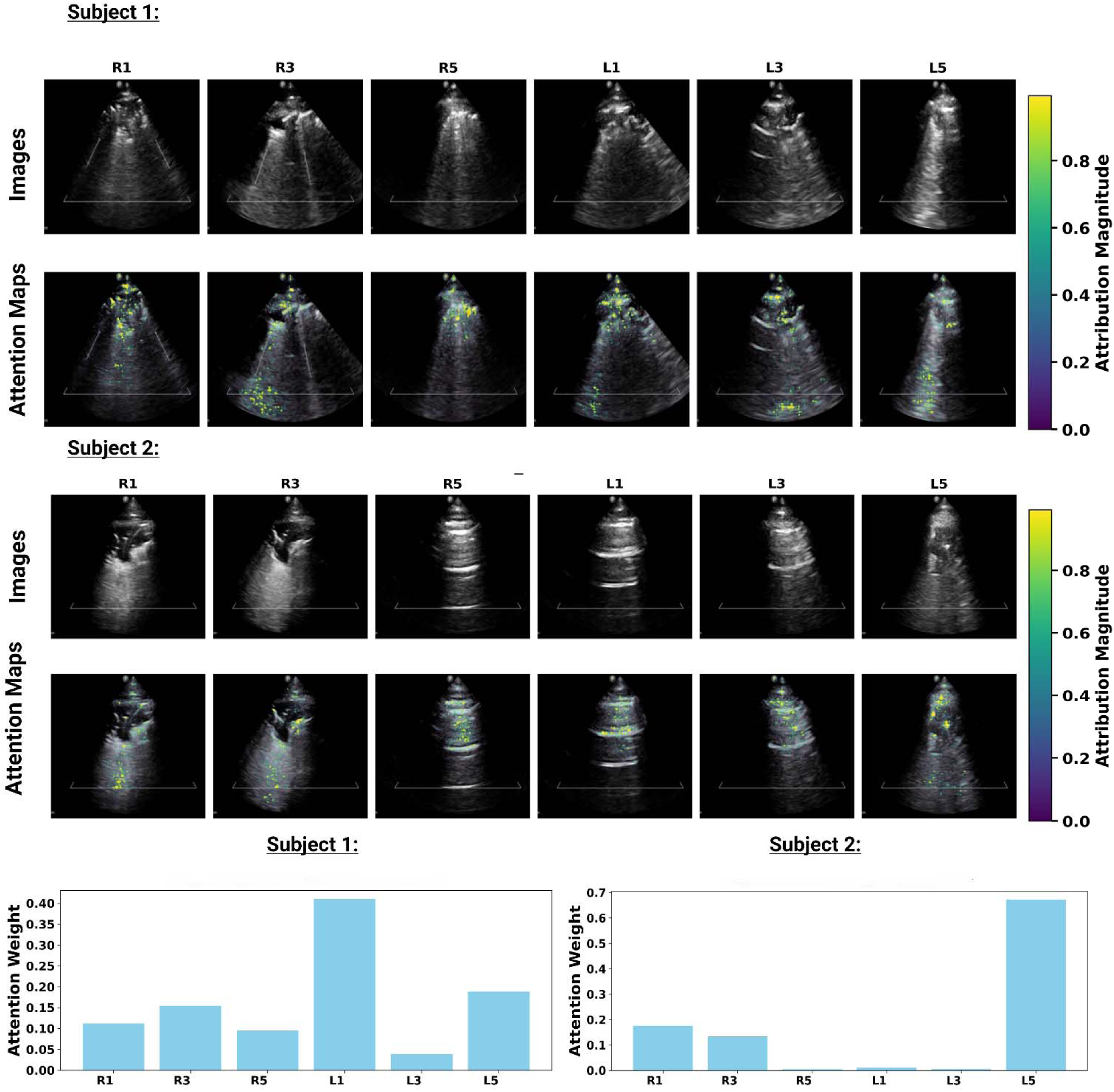
Attention weights and integrated-gradient maps for two participants with tuberculosis (6-zone ultrasound-only model). Native images with superimposed, pseudo-colored attribution magnitude, and bar plots of attention weight by zone. Alt text: Lung ultrasound images from six zones for two participants with tuberculosis, overlaid with color heat maps showing the image regions the model relied on. Bar charts beside the images show that the zones with the highest attention weights.

## IV. DISCUSSION

In this prospective Ugandan cohort, attention-based deep learning of a single still frame per lung zone discriminated microbiologically confirmed pulmonary TB, with test-set AUCs of 0.91–0.95. Models integrating LUS with routine clinical variables demonstrated high discrimination under both acquisition protocols, although differences in performance across models were not statistically significant. Point estimates for the combined models, including the streamlined 6-zone model using half the standard acquisition zones, exceeded the 2025 WHO minimum targets for a high-sensitivity screening test in a two-step algorithm (≥90% sensitivity, ≥60% specificity) (25) and the 2014 triage target (≥90% sensitivity at ≥70% specificity) (24). However, confidence intervals were wide, and their lower bounds fell below these targets for all models, so attainment remains to be confirmed with prespecified thresholds in larger cohorts. In illustrative cases, the zones the model weighted most heavily coincided with pathologic findings, and integrated-gradient maps localized these findings within individual images. Together, these findings suggest that SCAN-TB is a promising portable, radiation-free, sputum-independent triage approach requiring validation in larger studies.

This study adds to emerging evidence of strong AI-assisted LUS performance for TB triage, including direct TB prediction from selected images acquired through a 14-point sliding-scan protocol (5) and abnormality detection from streamlined ultrasound sweeps (6). We additionally performed participant-level TB classification across anatomically defined lung zones, directly compared clinical-only, ultrasound-only, and combined models, and included both symptomatic patients and household contacts, a target population for active case finding. We also explored an abbreviated 6-zone protocol selected using attention weights that retained discrimination, although the modest differences in weights do not definitely establish the optimal zone selection. The selected zones were all superior zones (anterior, lateral, and posterior), consistent with the upper-lobe predominance of pulmonary TB. This targeted approach may complement sweep-based protocols by standardizing regional sampling, although prospective comparison is needed to determine the relative efficiency and robustness of each.

The added value of combining imaging with clinical data is clinically plausible. LUS captures peripheral and pleural manifestations of pulmonary disease, whereas cough duration, weight loss, BMI, and general appearance reflect systemic disease and TB risk that ultrasound alone may not capture. Because these variables may be routinely collected during screening, integrating them adds little acquisition burden and may help distinguish TB from other conditions with overlapping sonographic findings. However, AUC differences between models were small (Table S8). The strong clinical-only performance likely reflects case mix, because TB-negative participants included asymptomatic household contacts who differed from TB+ participants largely in symptom duration.

This study has limitations to address before clinical implementation. First, models were developed and internally evaluated in a single Ugandan setting with one ultrasound device and trained operators. External validation across geographically and epidemiologically distinct cohorts, ultrasound platforms, operator groups, and patient populations will establish generalizability. Prospective implementation studies with intended end users, including non-specialist operators, should quantify acquisition failure rates, inter-operator variability, usability, and downstream effects on confirmatory testing and time to diagnosis. Second, TB+ participants were mostly index cases, whereas TB− participants were household contacts or clinic patients. This enrichment may overestimate performance in active case-finding populations, and with only four TB+ household contacts, none asymptomatic, performance in subclinical TB could not be assessed. Differences in clinical variable recording, completeness, and microbiological verification (Ultra alone versus Ultra plus culture) across recruitment groups may also introduce recruitment-related confounding and bias. Third, the test and validation sets were small and derived from a single random split. This precluded precise estimates across clinically important subgroups such as sex, HIV status, prior TB, disease severity, and smear status, as well as calibration, decision-curve, and net-benefit analyses. Consequently, whether the model performs equitably across these groups remains to be established. Fourth, a prespecified single-frame rule provides a standardized, reproducible input, but it omits temporal information such as respiratory dynamics and probe repositioning. Models using the middle frame remained discriminative but had lower AUCs than those using the first frame (0.85 vs 0.93 for ultrasound-only; 0.90 vs 0.95 for combined), indicating that performance varies by frame choice. Finally, performance was evaluated under a standardized research protocol and may differ in routine practice. At the same time, several strengths – including prospective design, microbiologically defined TB status, blinded and standardized LUS acquisition, participant-level separation of training and test data, and direct comparison of full and streamlined acquisition protocols - provide a foundation for future validation.

### Conclusion

Our clinically integrated deep learning framework for LUS showed promising internal performance for pulmonary TB triage, and a streamlined 6-zone acquisition protocol may enhance its feasibility at the point of care. If validated externally, this portable, radiation-free, sputum-independent approach could expand access to TB active case finding and improve targeting of confirmatory testing in resource-constrained settings.

## Author contributions

Conception and design: Y.L.X., A.M., J.E.; data acquisition: L.N., W.N., E.N., M.J., G.A.; image interpretation: K.G., W.N., E.N.; model development and analysis: A.S., R.D., A.M.; data curation and statistical analysis: E.J.J., R.S.; drafting: A.S., Y.L.X.; critical revision: all authors. Conflicts of interest: Conflicts of interest: Dr. Madabhushi is an equity holder in Picture Health, Elucid Bioimaging, and Inspirata Inc. He serves on the advisory board of Picture Health and currently consults for J&J. He has sponsored research agreements with AstraZeneca and Bristol Myers Squibb. His technology has been licensed to Picture Health and Elucid Bioimaging. None of these entities were involved in this work.

## Data availability

Data supporting the findings of this study will be deposited in a public repository upon publication. Until then, they are available from the corresponding author on reasonable request. Participants who declined storage and sharing of their data will be excluded from the shared dataset.

