## supplemental Materials for "Deep learning of point-of-care lung ultrasound and clinical data for pulmonary tuberculosis triage in Uganda"

**Supplementary figures and tables**

**Figure S1** — Anatomical lung ultrasound acquisition zones

**Table S1** — Clinical variable definitions and coding

**Table S2** — Participant characteristics by training/validation/test groups

**Figure S2** — Model architectures

**Table S3** — Image augmentation parameters

**Table S4** — Training parameters

**Figure S3** — Training and validation curves

**Table S5** — Baseline comparisons by TB status

**Table S6** — Lung-zone attention weights

**Table S7** — First-frame versus middle-frame sensitivity analysis

**Table S8** — Pairwise DeLong comparisons

**Figure S4** — Exploratory subgroup analyses

**Figure S5** — UMAP model representations

**Checklist** — TRIPOD+AI reporting checklist

**Figure S1.** Anatomical locations of the 12 lung ultrasound acquisition zones, shown from anterior and posterior views.

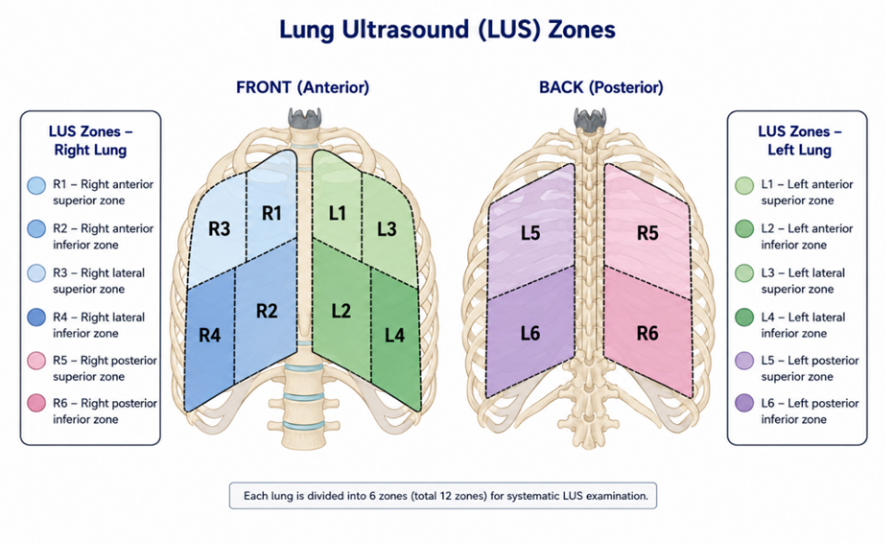

**Table S1.** Definitions and coding of clinical variables.

| **Variable** | **Source and definition** | **Coding in models** |
| --- | --- | --- |
| Age | Date of birth, or participant-estimated age if date of birth unknown | Continuous, years |
| Sex | Sex at birth | Binary (male, female) |
| Prior TB | Self-reported previous TB diagnosis | Binary (yes, no) |
| HIV status | Index cases and household contacts: documented result or study HIV testing. Participants with other respiratory conditions: documented history | Binary (positive, negative) |
| Diabetes | Self-reported or documented diagnosis of diabetes mellitus | Binary (yes, no) |
| Smoking duration | Total years of use of manufactured cigarettes, hand-rolled cigarettes, cigars, e-cigarettes, or hookah. Never smoked, or not applicable (adolescents), coded as 0 | Continuous, years |
| Cough duration | Reported duration of cough, recorded in days, weeks, or months. For participants with other respiratory conditions, dry and productive cough were recorded separately [longer duration used — confirm]. No cough coded as 0 | Continuous, weeks |
| Weight-loss duration | Reported duration of unintended weight loss, recorded in days, weeks, or months. None coded as 0 | Continuous, weeks |
| General appearance | Study staff rating: not ill; mildly ill; moderately ill; gravely ill (no participant was rated gravely ill) | Ordinal |
| Temperature | Digital thermometer at enrollment (temporal for participants with other respiratory conditions; [method] for index cases and household contacts) | Continuous, °C |
| Body mass index | Calculated from measured weight and height | Continuous, kg/m² |
| Household members | Total household members including the participant. Top-coded at 8 because the other respiratory condition case report form recorded 1 to ≥8 | Continuous, 1–8 |

**Table S2.** Participant characteristics by training/validation/test groups. Baseline characteristcs of the analyzed cohort (n = 316) by training (n = 223), internal validation (n = 20), and held-out test (n = 73) sets.

|  | **Missing** | **All (n=316)** | **Training (n=223)** | **Validation (n=20)** | **Test  (n=73)** | **p-value*** |
| --- | --- | --- | --- | --- | --- | --- |
| Patients N | 0 | 316 (100) | 223 (71) | 20 (6) | 73 (23) |  |
| Male, N (%) | 0 | 178 (56) | 123 (55) | 14 (70) | 41 (56) | 0.439 |
| Age (years); median (IQR) | 0 | 32 (25–43) | 32 (24–45) | 32 (26–37) | 30 (25–42) | 0.976 |
| BMI (kg/m²); median (IQR) | 0 | 20.6  (18.9–24.2) | 20.7  (18.9–24.5) | 19.4  (18.0–21.3) | 20.7  (19.2–23.4) | 0.115 |
| Temperature (°C); median (IQR) | 0 | 36.5  (36.2–36.8) | 36.5  (36.2–36.8) | 36.6  (36.2–36.8) | 36.4  (36.1–36.7) | 0.681 |
| **Comorbidities** |  |  |  |  |  |  |
| Prior TB history, N (%) | 0 | 17 (5) | 14 (6) | 0 (0) | 3 (4) | 0.423 |
| Diabetes, N (%) | 0 | 2 (1) | 2 (1) | 0 (0) | 0 (0) | 0.657 |
| HIV positive, N (%) | 0 | 64 (20) | 42 (19) | 2 (10) | 20 (27) | 0.143 |
| **Symptoms** |  |  |  |  |  |  |
| Cough duration (weeks); median (IQR) | 0 | 4 (1–12) | 4 (0–12) | 5 (3–8) | 4 (2–12) | 0.251 |
| Weight loss duration (weeks); median (IQR) | 0 | 0 (0–4) | 0 (0–4) | 4 (0–8) | 0 (0–8) | 0.038 |
| Ever-smoker, N (%) | 0 | 45 (14) | 27 (12) | 5 (25) | 13 (18) | 0.175 |
| **Signs** |  |  |  |  |  |  |
| General appearance | 0 |  |  |  |  | 0.994 |
| General appearance — Well, N (%) |  | 108 (34) | 77 (35) | 6 (30) | 25 (34) |  |
| General appearance — Moderate, N (%) |  | 150 (47) | 106 (48) | 10 (50) | 34 (47) |  |
| General appearance — Ill, N (%) |  | 58 (18) | 40 (18) | 4 (20) | 14 (19) |  |
| **Contact history** |  |  |  |  |  |  |
| Known TB contact, N (%)^a^ | 102 | 32 (15) | 17 (11) | 4 (27) | 11 (22) | 0.080 |
| Lived with TB patient, N (%)^a^ | 102 | 13 (6) | 7 (5) | 0 (0) | 6 (12) | 0.103 |
| Household members; median (IQR)^b^ | 0 | 4 (3–5) | 4 (3–6) | 4 (3–4) | 3 (3–5) | 0.174 |

Values are n (%) or median (interquartile range). TB = tuberculosis; BMI = body mass index. Contact-history items exclude participants with other respiratory conditions (n = 102); household size was top-coded at 8.

**Figure S2.** Model architecture. (A) Ultrasound-only image encoder and attention-based multiple-instance learning pooling. (B) Combined ultrasound–clinical model with feature fusion after attention pooling. M_i_ = ultrasound-only model; M_i+c_ = combined ultrasound and clinical-variable model.

**A.**

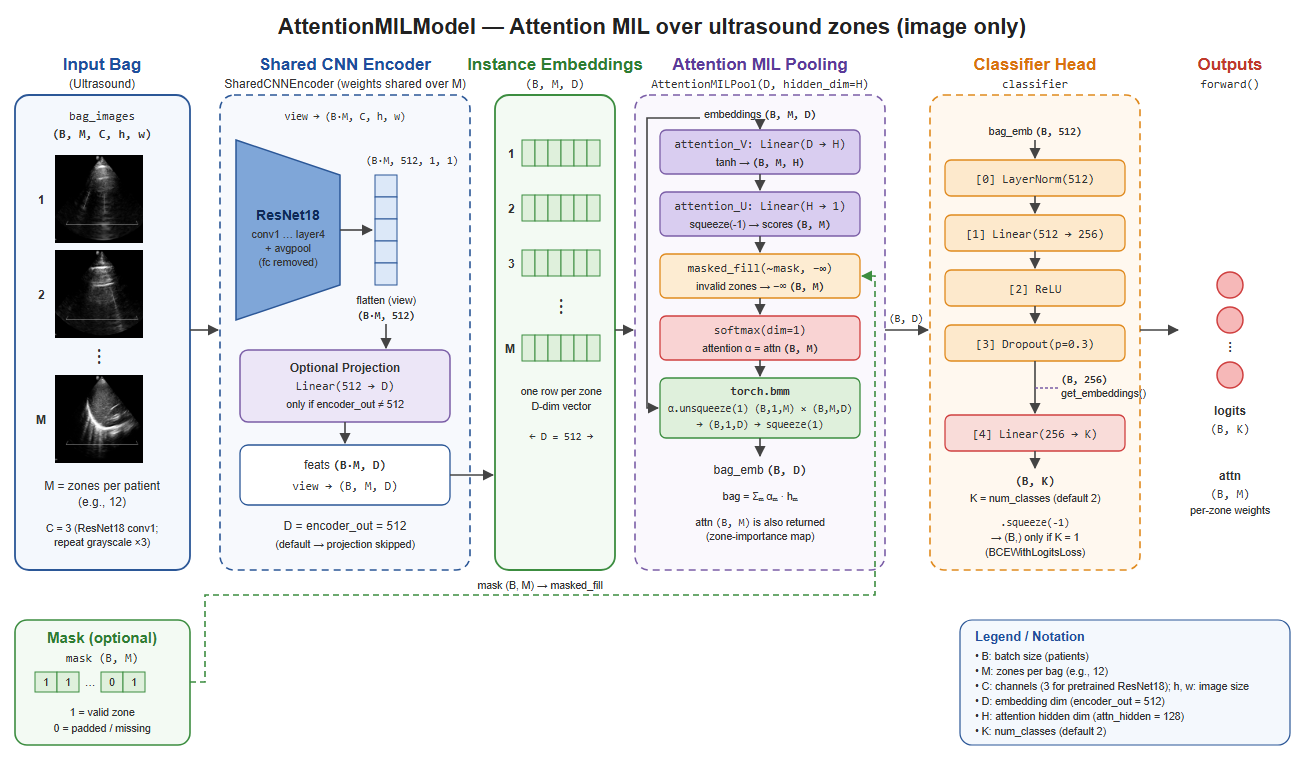

**B.**

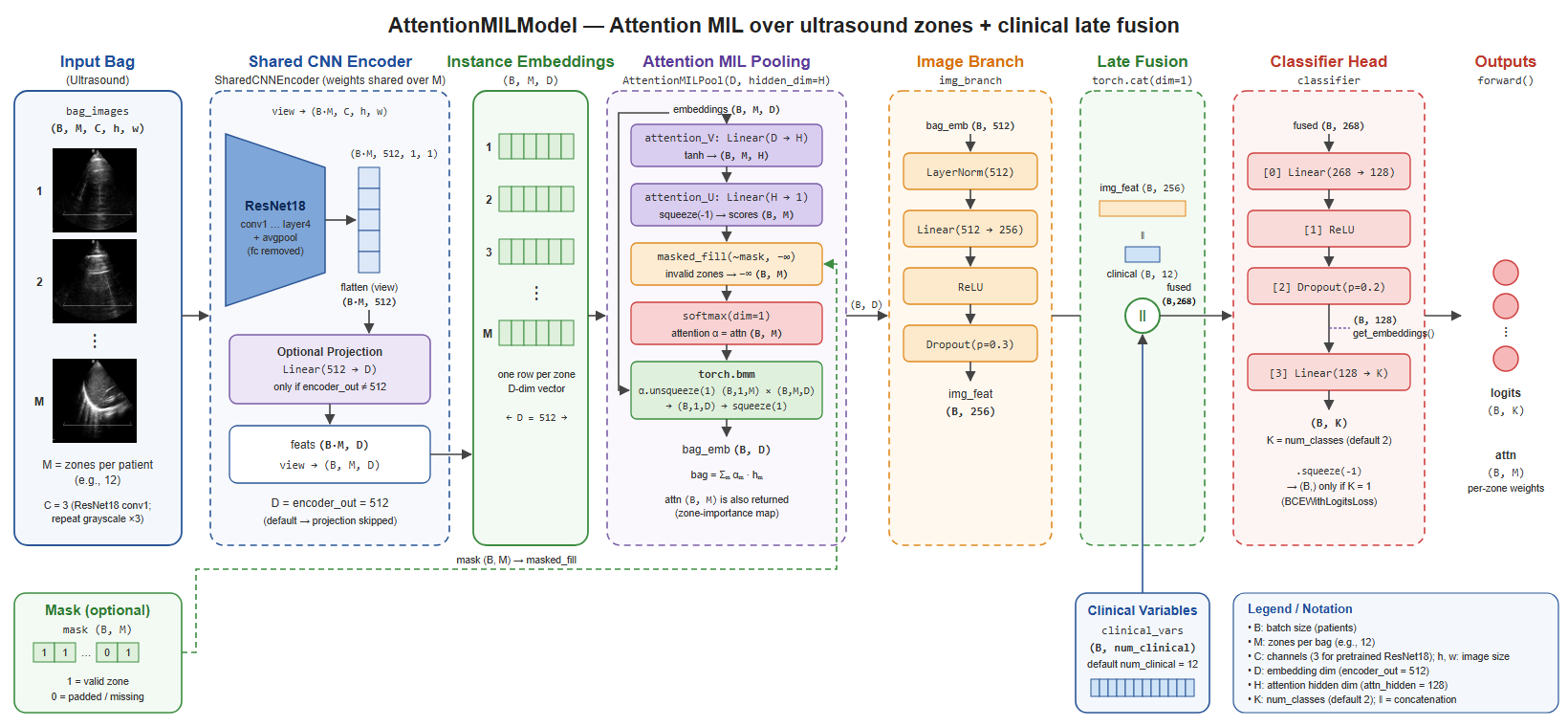

**Table S3.** Image augmentation transformations and parameters.

| **Transform** | **Parameters** |
| --- | --- |
| Resize | (224,224) |
| Horizontal Flip, Vertical Flip | P=0.5, P=0.5 |
| Random Brightness Contrast | P=0.2 |
| Hue Saturation | P=0.2 |
| Coarse Dropout | P=0.3, max holes=1, max height=50, max Width=50, min holes=1, min height=50, min width=50 |
| Rotate | Limit=30, p=0.5 |
| Shift Scale Rotate | Shift limit=0.1, scale limit=0.1, rotate limit=30, p=0.3 |
| Grid Distortion | P=0.5 |
| Gaussian Blur | Blur limit= (3,7), p=0.5 |
| Contrast-limited adaptive histogram equalization | P=0.5 |
| Z-normalize | Mean= [0.5,0.5,0.5]  Standard deviation= [0.5,0.5,0.5] |

**Table S4.** Deep learning training parameters.

**Training parameters**

Maximum Epochs=200

Loss function: Cross Entropy Loss

Optimizer: AdamW

Early stopping criterion with patience =35, min improvement in loss=1e-3

Weight Decay=1e-5

Learning rate=1e-4

Batch size=16

**Figure S3.** Training and validation loss and AUC during development of the A) Ultrasound only; M_i_ and B) Combined ultrasound and clinical variable; M_i+c_ models

**A.**

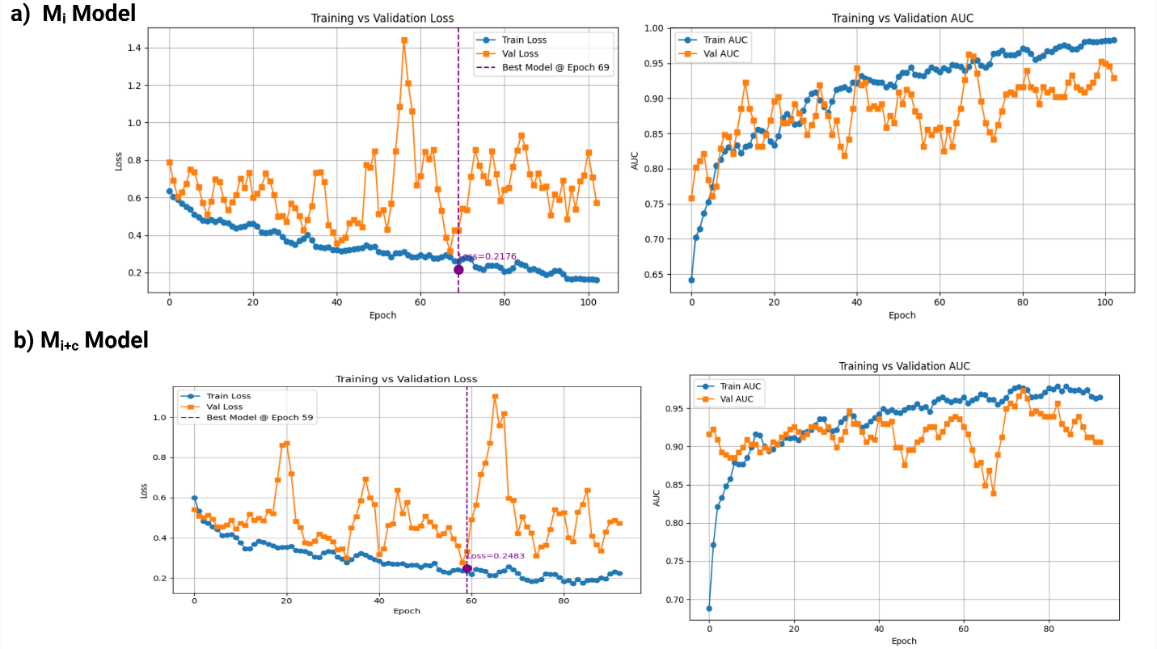

**B.**

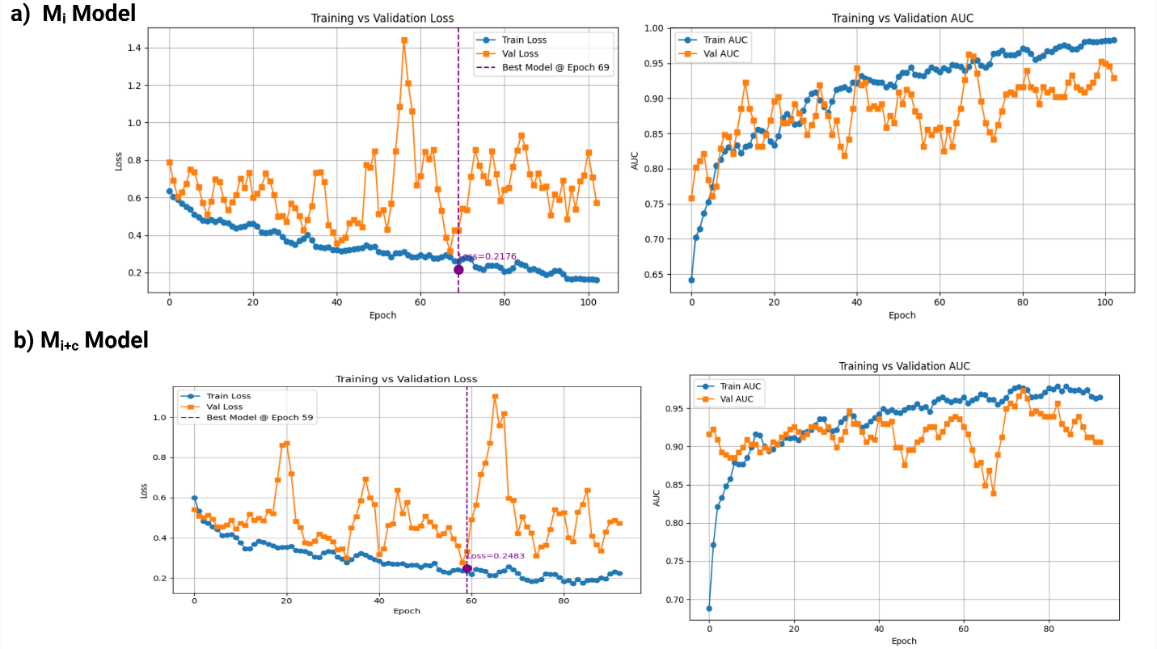

**Table S5.** Baseline clinical characteristics by tuberculosis status: reported between-group comparisons.

| **Characteristic** | **TB-positive vs TB-negative** | **Reported P value** |
| --- | --- | --- |
| Male | 61% vs 54% | 0.195 |
| Age, median (years) | 32 vs 32 | 0.648 |
| BMI, median (kg/m²) | 19.4 vs 22.4 | <0.001 |
| Temperature, median (°C) | 36.5 vs 36.4 | 0.091 |
| Prior TB history | 4% vs 6% | 0.487 |
| Diabetes | 1% vs 1% | 1.000 |
| HIV-positive | 19% vs 21% | 0.795 |
| Current or former smoking | 20.3% vs 10.6% | 0.017 |
| Cough duration, median (weeks) | 12 vs 2 | <0.001 |
| Weight-loss duration, median (weeks) | 4 vs 0 | <0.001 |
| General appearance (overall) | Three-category comparison | <0.001 |

Values and P values are reproduced from the main manuscript Table 1; no additional analysis was performed. BMI = body mass index; TB = tuberculosis.

**Table S6.** Mean attention weight by lung zone among the 79 tuberculosis-positive training participants. The six highest-ranked zones were selected for the abbreviated protocol.

| **Zone** | **Mean_Attention_TB_POS** |
| --- | --- |
| **R5** | 0.1086 |
| **L5** | 0.0955 |
| **L3** | 0.0884 |
| **R1** | 0.0830 |
| **R3** | 0.0815 |
| **L1** | 0.0805 |
| R6 | 0.0802 |
| L4 | 0.0800 |
| R2 | 0.0789 |
| L2 | 0.0778 |
| R4 | 0.0748 |
| L6 | 0.0744 |

**Table S7.** Sensitivity analysis using the middle frame of each lung ultrasound acquisition loop (held-out test set, n = 73). Mi = ultrasound-only model; Mi+c = combined ultrasound and clinical-variable model.

Values are AUC (95% CI). First-frame and middle-frame analyses used the same held-out test participants.

| **12-zone model** | **First-frame AUC (95% CI)** | **Middle-frame AUC (95% CI)** |
| --- | --- | --- |
| Ultrasound only | 0.93 (0.86–0.97) | 0.85 (0.74–0.94) |
| Ultrasound plus clinical variables | 0.95 (0.89–0.99) | 0.90 (0.82–0.97) |

**Table S8.** Pairwise DeLong comparisons of test-set AUCs.

| **Protocol** | **Comparison** | **P value** |
| --- | --- | --- |
| 12-zone | Ultrasound vs clinical | 0.39 |
|  | Combined vs clinical | 0.10 |
|  | Combined vs ultrasound | 0.50 |
| 6-zone | Ultrasound vs clinical | 0.62 |
|  | Combined vs clinical | 0.26 |
|  | Combined vs ultrasound | 0.57 |
| 12-zone vs 6-zone | Ultrasound | 0.56 |
|  | Combined | 0.45 |
| AUC = area under the receiver operating characteristic curve. Clinical = clinical-only model; ultrasound = ultrasound-only model; combined = combined ultrasound and clinical model. The clinical-only model is the same in both protocols. All comparisons used the same 73 test participants. P values are two-sided and not adjusted for multiple comparisons. | | |

**Figure S4.** Exploratory model accuracy by sex, prior tuberculosis history, and HIV status in the held-out test set. M_i_ = ultrasound-only model; M_i+c_ = combined ultrasound and clinical-variable model.

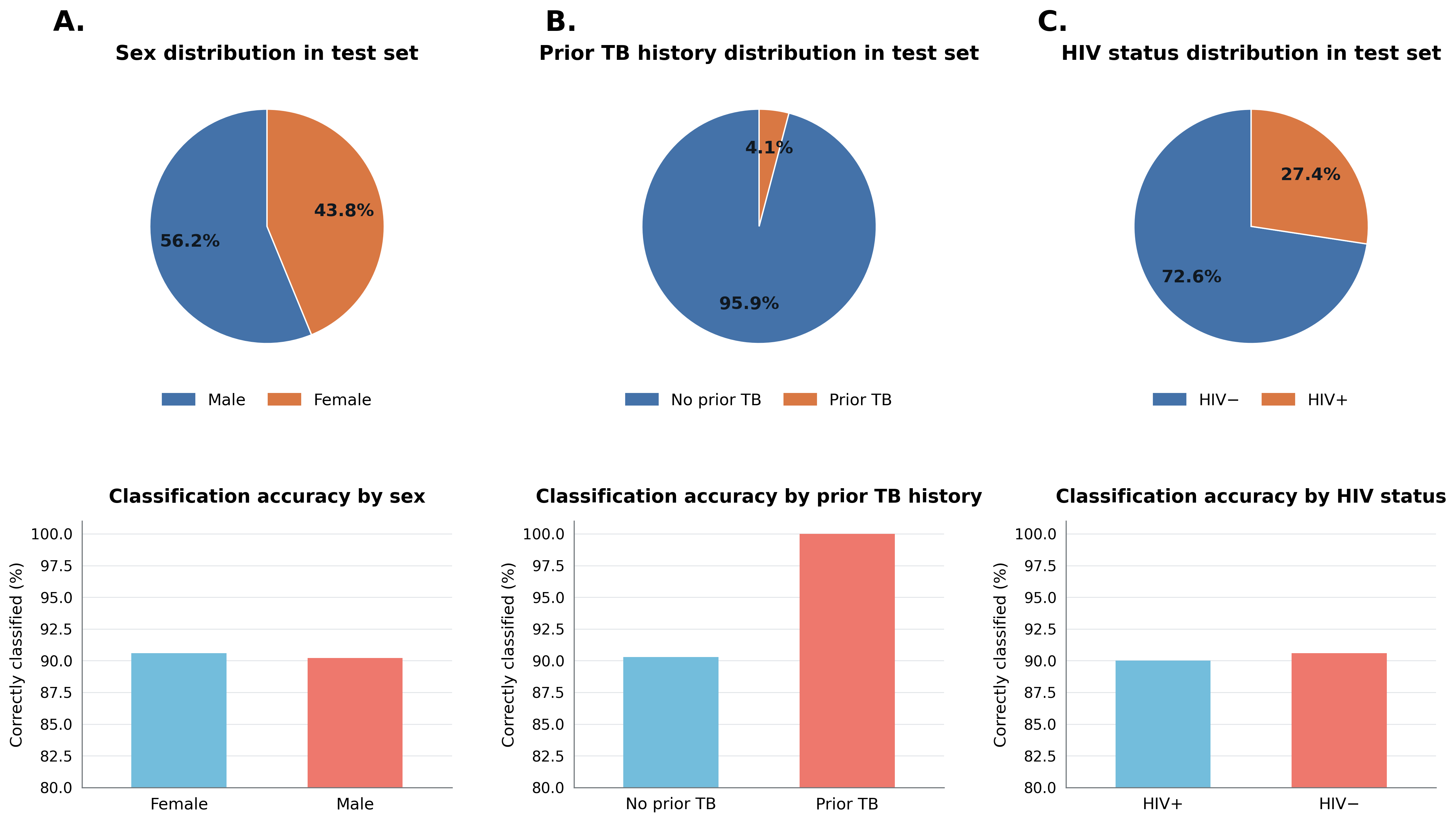

**Figure S5.** Exploratory UMAP visualizations of model representations using 6 zones. M_c_= clinical-variable only model; M_i_ = ultrasound-only model; M_i+c_ = combined ultrasound and clinical-variable model.

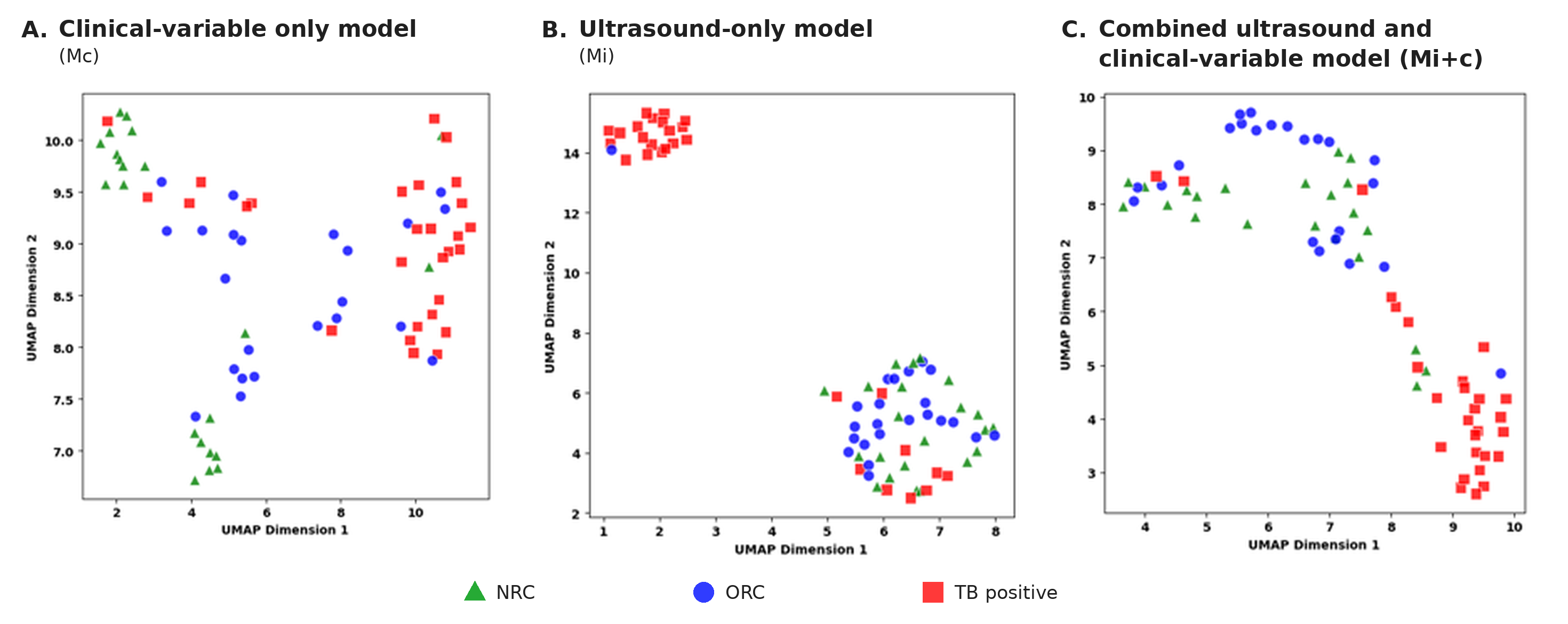

TRIPOD+AI reporting checklist

| **Section** | **Item** | **Checklist item (abbreviated)** | **Location in manuscript** | **Status** |
| --- | --- | --- | --- | --- |
| Title | 1 | Identify the study as developing or evaluating a prediction model, the target population, and the outcome | Title page | Reported |
| Abstract | 2 | Follow TRIPOD+AI for Abstracts | Abstract | Reported |
| Introduction: background | 3a | Healthcare context, rationale for the model, and existing models | Introduction, paragraphs 1–2 | Reported |
|  | 3b | Target population, intended purpose in the care pathway, and intended users | Introduction, paragraph 3; Discussion, paragraph 1 | Reported |
|  | 3c | Known health inequalities between sociodemographic groups | Not yet stated | Not reported |
| Introduction: objectives | 4 | Objectives, including whether the study develops and/or validates a model | Introduction, paragraphs 3–4 | Reported |
| Methods: data | 5a | Data sources, rationale, and representativeness | Study design and participants | Reported |
|  | 5b | Dates of participant accrual | Study design and participants (July 2023–March 2025) | Reported |
| Methods: participants | 6a | Setting, including number and location of centers | Study design and participants | Reported |
|  | 6b | Eligibility criteria | Study design and participants | Reported |
|  | 6c | Treatments received and how handled | Study design and participants (>15 days of TB treatment excluded) | Reported |
| Methods: data preparation | 7 | Preprocessing and quality checking, including consistency across groups | Deep learning ultrasound and combined models; Table S2 | Reported |
| Methods: outcome | 8a | Outcome definition, timing, rationale, and consistency of assessment across groups | Reference standard and participant classification | Reported |
|  | 8b | Qualifications of outcome assessors if assessment is subjective | Reference standard and participant classification | Reported |
|  | 8c | Blinding of outcome assessment | Lung ultrasound acquisition | Reported |
| Methods: predictors | 9a | Choice of predictors and any pre-selection | Clinical variables | Reported |
|  | 9b | Definition and measurement of predictors, and blinding | Clinical variables; Table S8; Lung ultrasound acquisition | Reported |
|  | 9c | Qualifications of predictor assessors | Lung ultrasound acquisition (two trained, certified technicians); Table S8 | Reported |
| Methods: sample size | 10 | How study size was determined and justified | Sample size | Reported |
| Methods: missing data | 11 | Handling of missing data and reasons for omissions | Lung ultrasound acquisition; Table 1 | Partially reported (partially missing zones pending; comment c15) |
| Methods: analytical methods | 12a | How data were used and partitioned | Data partitioning | Partially reported (random seed and checkpoint criterion pending) |
|  | 12b | Predictor handling (rescaling, standardization) | Clinical variables; Table S8 | Reported |
|  | 12c | Model type, building steps, hyperparameter tuning, and internal validation | Clinical variables; Deep learning ultrasound and combined models; Table S3 | Reported |
|  | 12d | Heterogeneity across clusters (e.g., sites) | Statistical analysis | Not reported due to small number of test participants per site. |
|  | 12e | Performance measures and model comparisons | Statistical analysis | Reported |
|  | 12f | Model updating or recalibration | — | Not applicable |
|  | 12g | How predictions were calculated | Not yet stated | Not reported (code availability) |
| Methods: class imbalance | 13 | Class imbalance methods, if used | Deep learning ultrasound and combined models;  No resampling or class weighting | Reported |
| Methods: fairness | 14 | Approaches to address model fairness | Statistical analysis (exploratory subgroup accuracy) | Partially reported |
| Methods: model output | 15 | Model output and how thresholds were identified | Statistical analysis | Reported |
| Methods: training versus evaluation | 16 | Differences between development and evaluation data | Data partitioning; Table S1 | Reported |
| Methods: ethics | 17 | Ethics approval and informed consent | Ethics statement (Mulago Hospital REC 2349; Rutgers IRB Pro2020003026) | Reported |
| Open science | 18a | Funding and funder role | Title page: Funding | Reported (PLOS: move to submission form) |
|  | 18b | Conflicts of interest | Title page: Conflicts of interest | Not reported (placeholder) |
|  | 18c | Protocol access | Online Supplement | Not reported (protocol to be added) |
|  | 18d | Study registration | Online Supplement: This observational study was not prospectively registered. | Reported |
|  | 18e | Data availability | Not yet stated | Not reported |
|  | 18f | Code availability | Not yet stated | Not reported |
| Patient and public involvement | 19 | Patient and public involvement, or a statement of none | Online Supplement: Patients and the public were not involved in the design, conduct, reporting, or dissemination of this research. | Reported |
| Results: participants | 20a | Participant flow, including numbers with and without the outcome | Baseline characteristics; Figure 2 | Reported |
|  | 20b | Characteristics, key dates, predictors, outcome events, and missing data | Table 1; Table S1 | Reported |
|  | 20c | Comparison of development and evaluation data | Table S1 | Reported |
| Results: model development | 21 | Participants and outcome events in each analysis | Baseline characteristics; Selection of the 6-zone protocol | Reported |
| Results: model specification | 22 | Full model details or access for third parties | Methods; Tables S2–S3; Figure S2 | Partially reported |
| Results: model performance | 23a | Performance with CIs, including key subgroups | Ultrasound-only and combined models; Table 2; Figure 4; Subgroup analysis; Figure S4 | Partially reported (DeLong comparisons and subgroup CIs pending) |
|  | 23b | Heterogeneity across clusters | — | Not reported due to small number of participants per site/cluster (see item 12d) |
| Results: model updating | 24 | Results of model updating | — | Not applicable |
| Discussion: interpretation | 25 | Overall interpretation, including fairness, in context of prior studies | Discussion, paragraphs 1–4 | Reported |
| Discussion: limitations | 26 | Limitations and effects on bias, uncertainty, and generalizability | Discussion, limitations paragraph | Reported |
| Discussion: usability | 27a | Handling of poor-quality or unavailable input data in practice | Not yet stated | Not reported |
|  | 27b | User interaction and required expertise | Discussion, limitations paragraph | Reported |
|  | 27c | Next steps for future research | Discussion, limitations paragraph; Conclusion | Reported |
